# Race, ethnicity, and risk for colonization and infection with key bacterial pathogens: a scoping review

**DOI:** 10.1101/2024.04.24.24306289

**Authors:** Esther E. Avendano, Sarah Addison Blackmon, Nanguneri Nirmala, Courtney W. Chan, Rebecca A. Morin, Sweta Balaji, Lily McNulty, Samson Alemu Argaw, Shira Doron, Maya L. Nadimpalli

## Abstract

**Background:** Racial and ethnic disparities in infectious disease burden have been reported in the USA and globally, most recently for COVID-19. It remains unclear whether such disparities also exist for priority bacterial pathogens that are increasingly antimicrobial-resistant. We conducted a scoping review to summarize published studies that report on colonization or community-acquired infection with pathogens among different races and ethnicities.

**Methods:** We conducted an electronic literature search of MEDLINE®, Daily, Global Health, Embase, Cochrane Central, and Web of Science from inception to January 2022 for eligible observational studies. Abstracts and full-text publications were screened in duplicate for studies that reported data for race or ethnicity for at least one of the pathogens of interest.

**Results:** Fifty-four observational studies in 59 publications met our inclusion criteria. Studies reported results for *Staphylococcus aureus* (n=56), *Escherichia coli* (n=8)*, Pseudomonas aeruginosa* (n=2), Enterobacterales (n=1), *Enterococcus faecium* (n=1), and *Klebsiella pneumoniae* (n=1), and were conducted in the USA (n=42), Israel (n=5), New Zealand (n=4), Australia (n=2), and Brazil (n=1). USA studies most often examined Black and Hispanic minority groups and regularly reported a higher risk of these pathogens in Black persons and mixed results for Hispanic persons. Ethnic minority groups were often reported to be at a higher risk in other countries.

**Conclusion:** Sufficient evidence was identified to justify systematic reviews and meta-analyses evaluating the relationship between race, ethnicity, and community-acquired *S. aureus* and *E. coli,* although data were rare for other pathogens. We recommend that future studies clarify whether race and ethnicity data are self-reported, collect race and ethnicity data in conjunction with the social determinants of health, and make a concerted effort to include non-English speakers and Indigenous populations from the Americas, when possible.

## INTRODUCTION

Racial and ethnic disparities in communicable disease risk have widely been reported in the USA and other countries.^1,2^ Recently, stark disparities in COVID-19 hospitalizations and deaths among racial and ethnic minority groups across several countries have drawn renewed attention to this challenge. Antimicrobial-resistant (AMR) bacterial infections are an ongoing threat to global public health.^3,4^ AMR infections are often acquired outside of healthcare settings, *i.e.*, in the community, with individuals who are colonized or carrying these organisms in the gut, on the skin, in the nares, or on other mucosal surfaces often at higher risk for subsequent infection with the same organism.^5,6^ Compared to infections with susceptible pathogens, infections with AMR pathogens lead to significantly longer hospital stays, increased healthcare-associated costs, and higher risks of death.^7^

Several studies have documented differences in pathogen colonization and infection risks between distinct racial and ethnic groups.^8–10^ However, to date, this information has not been synthesized for priority bacterial pathogens that are increasingly AMR, including *Enterococcus faecium*, *Staphylococcus aureus*, *Klebsiella pneumoniae*, *Acinetobacter baumannii*, *Pseudomonas aeruginosa*, *Enterobacter* species, and *Escherichia coli*. There is no biological basis for racial or ethnic differences in rates of colonization or infection with priority bacterial pathogens; nevertheless, such differences likely exist because race and ethnicity are closely intertwined with individuals’ socioeconomic status (SES) and the social determinants of health (SDOH) more broadly, at least in part due to structural racism.^11^ While using more specific indicators than dichotomous race or ethnic status would be preferable to improve generalizability for patient care,^12^ variables related to the SDOH can be challenging to measure and have not traditionally been collected in many healthcare settings.^12^ Although there is increasing momentum for these data to be collected, collection remains inconsistent in many healthcare settings due to lack of standardized guidance on how to collect and interpret SDOH data,^13^ as well as constraints on clinicians’ time.^14^ Thus, compiling global evidence for the association between an individual’s racial or ethnic status and their risk of colonization or infection with priority bacterial pathogens is valuable for summarizing current evidence, assessing gaps, and guiding more nuanced data collection in the future.

Scoping reviews are used to compile and descriptively summarize current published evidence on a scientific topic, as well as to identify gaps where additional research could be beneficial. Our specific objective in this scoping review was to compile and summarize existing evidence for an association between individuals’ racial or ethnic background and their risk of colonization or infection with select community-acquired bacterial pathogens that are increasingly AMR.

## METHODS

### Search strategy

The search strategy for this scoping review was constructed to support the current study as well as a scoping review of the evidence for the association between SES and disparities in community-acquired colonization/infection with the same bacteria.^15^ A comprehensive scientific literature search of MEDLINE (Ovid), MEDLINE Epub Ahead of Print, In-Process, In-Data-Review & Other Non-Indexed Citations, and Daily (Ovid), Global Health (Ovid), Embase (Elsevier), Cochrane Database of Systematic Reviews (Wiley), Cochrane Central Register of Controlled Trials (Wiley), and Web of Science Core Collection was conducted for studies that reported race, ethnicity, and at least one of the pathogens of interest. Search strategies were designed as a joint effort between team members, experts, and a librarian using a combination of controlled vocabulary and free-text keywords. All searches were based on an initial MEDLINE search and utilized MeSH terminology and related keywords for the following concepts: Community-Acquired Infections, Outpatients, Ambulatory Care, Socioeconomic Factors, Health Status Disparities, Healthcare Disparities, Continental Population Groups, Ethnic Groups, Gram-Negative Bacteria, and individual ESKAPE pathogens. The MEDLINE strategy (**Supplementary Table S1**) was translated to each of the listed databases by Reference Manager, and all databases were searched from inception through January 2022, except for MEDLINE Epub Ahead of Print, In-Process, In-Data-Review & Other Non-Indexed Citations and Daily for which the search covered 2017 through 10 January 2021. References were collected and deduplicated using Endnote X9, prior to export to Covidence for screening and management.^16^

### Eligibility Criteria

We included peer-reviewed publications of any study design, except case-control or case-series, that reported data on race or ethnicity for at least one of the following pathogens: *Enterococcus faecium*, *Staphylococcus aureus*, *Klebsiella pneumoniae*, *Acinetobacter baumannii*, *Pseudomonas aeruginosa*, *Enterobacter* spp., or *Escherichia coli*, or for Enterobacterales (formerly called Enterobacteriaceae), an order which comprises several of our species of interest, *including E. coli*, *K. pneumonie*, and *Enterobacter* spp. We did not limit our inclusion criteria by age, country, or publication language and we included both infection and colonization (**Supplementary Table S2**). We excluded studies that reported infections with mixed pathogens if less than 50% of the pathogens were of interest unless subgroup data were reported for at least one pathogen of interest. We only included studies that reported outpatient or community-based data or specified that the patient’s pathogen of interest was community-acquired. Studies that did not report outpatient or community data and which defined community acquisition based only on phenotype (e.g., susceptibility to gentamicin for *S. aureu*s) or sequence type (e.g., USA300 for *S. aureu*s), were excluded. We also excluded studies that reported hospital-acquired pathogens in the comparator group. In studies reporting on colonization, we excluded studies that compared persistent versus cleared colonization.

We note that interpretations of race, ethnicity, and “minority” groups vary by country and their historical contexts. We used authors’ classifications of race and ethnicity whenever possible; for example, we included Hispanic as a race when authors reported it as such, even though current USA guidelines categorize Hispanic/Latino as an ethnicity. In papers from Israel, we categorized Bedouins, non-Bedouin Arabs, and Jewish persons as ethnicities, even though Bedouins and Arabs are a tribal or religious minority rather than a true racial or ethnic minority, and Jewish persons are of multiple races. We included these Israeli studies because they intended to examine the effect of minority status on colonization/infection with the pathogens of interest. We did not use the terms “White”, “Caucasian”, or “European” interchangeably in our data extraction or synthesis; rather, we used terms used by each study’s authors.

### Study Selection, Data Extraction, and Synthesis

Titles and abstracts of citations identified from the literature searches were screened in duplicate in Covidence using the predefined inclusion/exclusion criteria created in conjunction between team members and experts (**Supplementary Table S2**).^16^ Any conflicts were resolved during team meetings, and full-text articles for any abstracts that met the inclusion criteria were retrieved and independently screened by two team members. A customized extraction form was created in Covidence by experienced team members to capture all relevant study data, including study design features, study definition of community-acquired, study population characteristics including race, ethnicity, exposure and comparator, outcomes of interest, and directionality of results. All team members piloted the extraction form to refine and capture all pertinent data. After the initial training, each study was independently extracted by two team members, and any discrepancies between the two were resolved by a third team member. We extracted all comparisons between races or ethnicities and outcomes reported in each paper regardless of whether the authors conducted a statistical test. Extracted data from all included studies are summarized across common parameters in narrative form, tables, and figures using R and Microsoft Excel 2021. This study followed the PRISMA checklist for scoping reviews (**Supplementary Material**).

## RESULTS

Our literature search, which supported the present study as well as a separate review of the evidence for the association between SES and disparities in community-acquired colonization/infection,^15^ identified 1039 citations that potentially reported data for race, ethnicity, or SES for patients colonized or infected with at least one of the pathogens of interest. Of these, 388 abstracts met the eligibility criteria and proceeded to full-text review (**Supplementary Figure S1**) of which 59 publications (**Supplementary Table S3**) were found to meet all inclusion criteria for race and ethnicity.^3,8–10,17–71^ Four cohort studies were described in 2-3 separate publications each, meaning 54 distinct studies were identified in total for the present study.

### Study characteristics

Among the 54 included studies,^3,8–10,17–71^ 42 (78%) described infections and 12 (22%) described colonization with at least one of the pathogens of interest (**Table 1**).^17,19,23,31,38,42,47,48,50,53,55–57,59^ Most studies were from the USA (n=42),^3,8–10,17,20–22,25,26,26,28–33,35–38,42,43,45,46,48–53,55–65,69–71^ although studies from Israel (n=5),^19,34,39,41,44,54^ New Zealand (n=4),^18,47,66–68^ Australia (n=2),^24,40^ and Brazil (n=1)^23^ were also identified (**Figure 1**). Studies of infection most often included patients with skin and soft tissue infection (SSTI) caused by *S. aureus* or urinary tract infection (UTI) caused by *E. coli* or other Enterobacterales, while other infections like community-acquired pneumonia (CAP) were described more rarely. Study definitions of community-acquired infection included no hospitalization within the last 30 days or one year or samples collected either at admission or up to 72 hours after admission. For studies that reported on colonization, samples were typically collected from the skin or nares.

**Figure 1.**
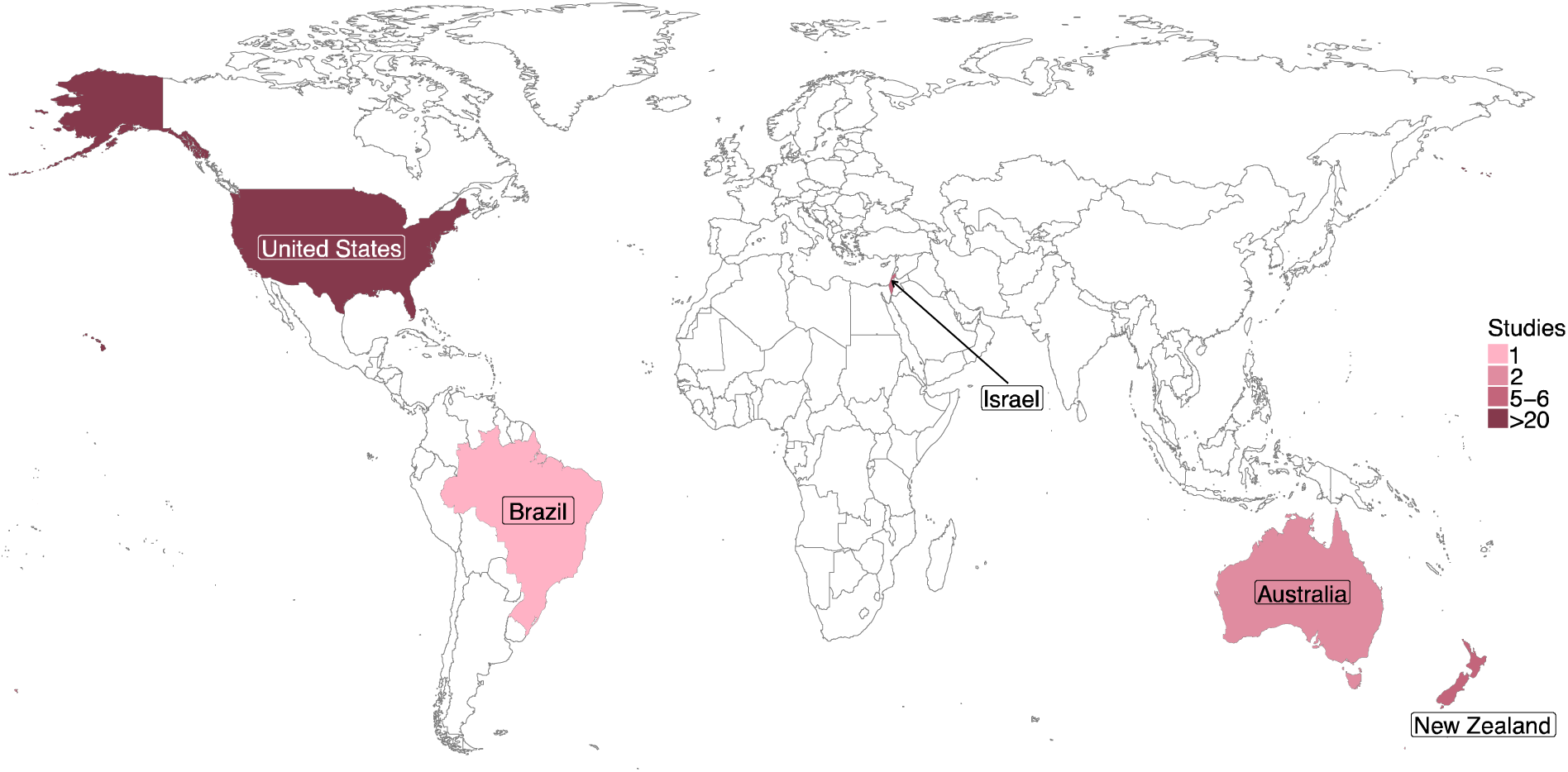
Study location for 54 studies describing individuals’ risk of colonization or infection with priority pathogens by their racial or ethnic identity.

**Table 1.** Characteristics of all included studies.

|  | **Total (%)** |
| --- | --- |
| **N studies** | 54 in 59 publications |
| **Age group** |  |
| Pediatric only | 22 (40.7%) |
| Adults only | 17 (31.4%) |
| Mixed age groups | 15 (27.8%) |
| **N analyzed range** | 33- 1,904,807 |
| **Study duration** | 4 months- 9 years |
| **% Male range (N=43)** | 5.4- 98.1% |
| Women only | 4 ( 7.4%) |
| **Study country** |  |
| Australia | 2 ( 3.7%) |
| Brazil | 1 ( 1.8%) |
| Israel | 5 ( 9.3%) |
| New Zealand | 4 ( 7.4%) |
| USA | 42 (77.8%) |
| **Outcome type** |  |
| Colonization | 12 (22.2%) |
| Infection | 42 (77.8%) |
| **Recruitment setting** |  |
| Inpatient | 21 (38.8%) |
| Outpatients | 14 (25.9%) |
| Both Inpatients and Outpatients | 13 (24.1%) |
| Community | 5 ( 9.3%) |
| Jail | 1 ( 1.9%) |
| **Pathogen** |  |
| MRSA | 37 (68.5%) |
| Other *Staphylococcus aureus* | 19 (35.2%) |
| *Enterococcus faecium* | 1 ( 1.9%) |
| *Escherichia coli* | 8 (14.8%) |
| *Pseudomonas aeruginosa* | 2 ( 3.7%) |
| *Klebsiella pneumoniae* | 1 ( 1.9%) |
| Enterobacterales | 1 ( 1.9%) |
| **Funding type (N=59)** |  |
| Academia only | 2 ( 3.3%) |
| Federal only | 16 (27.1%) |
| Industry only | 2 ( 3.3%) |
| Mixed | 13 (22.0%) |
| None | 9 (15.3%) |
| Not reported | 17 (28.8%) |

Note: MRSA=Methicillin-resistant *S. aureus*.

### Race

#### Black vs. White persons

Thirty-two observational studies in 35 publications compared Black persons to White persons (**Table** 2, Figure 2).^8–10,17,20–23,27,27,29,31–33,35–38,42,43,45,46,48–51,55,60–62,64,69–71^ Twenty-five observational studies in 26 publications reported on Methicillin-resistant Staphylococcus aureus (MRSA) or *S. aureus*,^9,10,20–22,26,29–33,35,37,38,42,43,45,49–51,55,58,59,62,64,69^ four on *E. coli*,^27,60,61,70^ one on Enterobacterales,71 one on SSTI caused by varied pathogens,^27^ and one on UTI caused by varied pathogens.^46^ Most studies reported Black persons to be at higher risk of MRSA colonization or infection, at either a lower or equivalent risk of *S. aureus* colonization and infection, and at either a higher or no difference in risk for infection with other pathogens.

**Figure 2.**
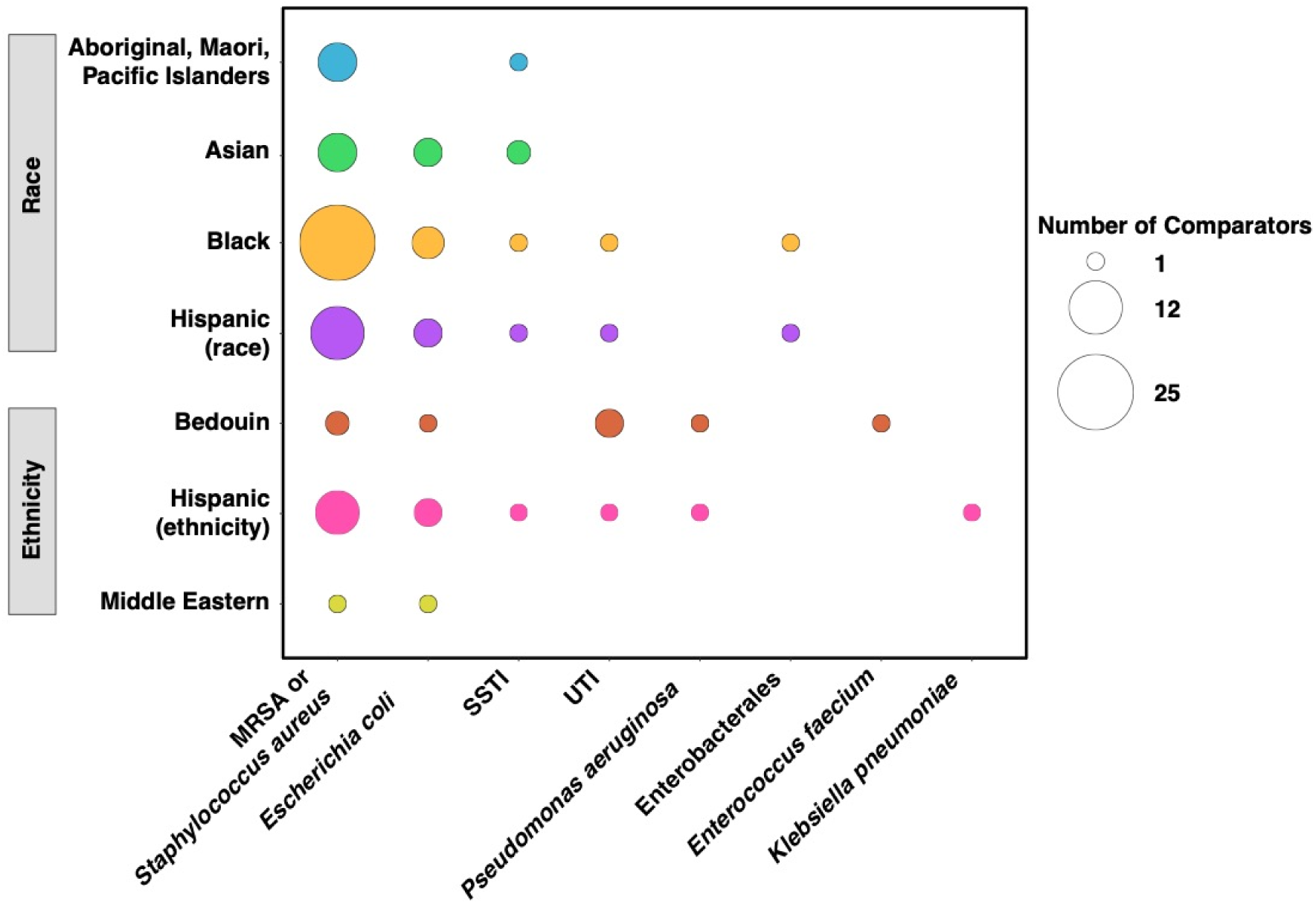
Frequency of reports of priority bacterial pathogens or associated infections by racial or ethnic minority groups vs majority group, among 54 studies included in this scoping review. Note: MRSA=Methicillin-resistant *S. aureus*. UTI=Urinary tract infection; SSTI=skin and soft. tissue infection.

Fifteen studies reported that Black individuals were at a higher risk of MRSA infection or colonization than White individuals.^9,10,20,22,31,37,38,43,45,49,50,55,62,64,69^ A multi-city prospective cohort study in the USA reported a higher risk of MRSA infection in Black to White persons in a southeastern urban center (Atlanta), but a lower risk in a mid-Atlantic city (Baltimore).^37^ Black adult outpatients in South Texas were reported to be at a higher risk of *S. aureus* SSTI that were non-susceptible to three or more antibiotic classes, i.e., multidrug-resistant (MDR) versus non-MDR *S. aureus* SSTI in one study, compared to participants who were neither Black nor Hispanic.^51^ One study conducted in Brazil with a total of 300 participants reported a more than five times higher risk of MRSA colonization vs. no MRSA colonization among Black participants than those who were not part of the Black minority.^23^ Five observational studies in six publications did not find a significant difference in MRSA infection or colonization between races,^21,26,32,33,42,50^ and four did not statistically compare MRSA among races.^29,35,42,59^

Some studies also examined differences in MSSA or non-resistant *S. aureus* among races, with mixed results. Two studies in California and Texas reported no significant difference in infection between Black and White participants,^8,62^ two in Georgia and Ohio reported no difference between Black persons and other races in infection or colonization rates,^33,48^ and one reported a lower risk of colonization in mixed-age Black participants compared to White participants.^42^

Ray 2013 also reported a lower risk of SSTI vs no SSTI in a mixed-age population of Black persons in metro Atlanta compared to the White population,^62^ while Casey 2013 did not find a difference between Black and White patients in central Pennsylvania with SSTI compared to matched controls.^26^

Enterobacterales infections in Black persons were examined in six studies. Among four studies reporting *E. coli* infections,^27,60,61,70^ two studies in San Francisco and Philadelphia reported no significant difference between Black and White participants,^60,61^ and two did not investigate differences between races for extended-spectrum beta-lactamase (EBSL)-producing-vs non-ESBL-producing *E. coli* or MDR vs non-MDR *E. coli* infections.^27,70^ One retrospective cohort study that included 40,137 participants from 175 US hospitals found Black adults to be at a higher risk than White adults of carbapenem-resistant Enterobacterales (CRE) vs susceptible Enterobacterales infections,^71^ while one study that reported on pregnant women with and without antepartum pyelonephritis caused by *E. coli* or *Enterobacter* spp. in Dallas, Texas did not find a difference between races.^46^

#### Hispanic (reported as race) vs. White persons

Seventeen studies reported Hispanic as a racial group (**Table 3**), all of which were conducted in the US.^9,10,21,25,26,32,33,45,46,49,51,59,60,62,69–71^ Conflicting patterns were reported regarding Hispanic persons’ risk of MRSA or *S. aureus* colonization/infection in 12 studies, Enterobacterales in five, and SSTI and UTI each in one study.

**Table 2:** Characteristics of studies that included Black persons.

|  | **Total (%)** |
| --- | --- |
| **N studies** | 32 in 35 publications |
| **Age group** |  |
| Pediatric only | 10 (31.3%) |
| Adults only | 13 (40.6%) |
| Mixed age groups | 9 (28.1%) |
| **N analyzed range** | 33- 1904807 |
| **Study duration** | 56 days- 11 years |
| **% Male range (N=25)** | 12.5- 93% |
| Women only | 4 (12.5%) |
| **Study country** |  |
| Australia | 0 (0%) |
| Brazil | 1 ( 3.1%) |
| Israel | 0 (0%) |
| New Zealand | 0 (0%) |
| USA | 31 (96.9%) |
| **Outcome type** |  |
| Colonization | 7 (21.9%) |
| Infection | 25 (78.1%) |
| **Recruitment setting** |  |
| Inpatient | 11 (34.4%) |
| Outpatients | 10 (31.3%) |
| Both Inpatients and Outpatients | 9 (28.1%) |
| Community | 1 ( 3.1%) |
| Jail | 1 ( 3.1%) |
| **Pathogen** |  |
| MRSA or *Staphylococcus aureus* | 25 (78.1%) |
| *Enterococcus faecium* | 0 (0%) |
| *Escherichia coli* | 4 (12.5%) |
| *Pseudomonas aeruginosa* | 0 (0%) |
| *Klebsiella pneumoniae* | 0 (0%) |
| Enterobacterales | 1 ( 3.1%) |
| SSTI | 1 ( 3.1%) |
| UTI | 1 ( 3.1%) |
| **Funding type** |  |
| Academia only | 1 ( 3.1%) |
| Federal only | 9 (28.1%) |
| Industry only | 2 ( 6.3%) |
| Mixed | 9 (28.1%) |
| None | 4 (12.5%) |
| Not reported | 7 (21.9%) |
| **Reported directionality of effect measure, compared to White majority** |  |
| Higher risk | 18 (56.3%) |
| No significant difference | 12 (37.5%) |
| Lower risk | 3 ( 9.4%) |

Note: MRSA=Methicillin-resistant *S. aureus*. UTI=Urinary tract infection; SSTI=skin and soft tissue infection.

**Table 3.** Characteristics of studies that included Hispanic persons as a race.

|  | **Total (%)** |
| --- | --- |
| **N studies** | 17 |
| **Age group** |  |
| Pediatric only | 6 (35.3%) |
| Adults only | 7 (41.2%) |
| Mixed age groups | 4 (23.5%) |
| **N analyzed range** | 33- 376262 |
| **Study duration** | 6 months- 10 years |
| **% Male range (N=14)** | 5.4- 80% |
| Women only | 2 (11.8%) |
| **Study country** |  |
| Australia | 0 (0%) |
| Brazil | 0 (0%) |
| Israel | 0 (0%) |
| New Zealand | 0 (0%) |
| USA | 17 (100%) |
| **Outcome type** |  |
| Colonization | 1 (5.9%) |
| Infection | 16 (94.1%) |
| **Recruitment setting** |  |
| Inpatient | 7 (41.2%) |
| Outpatients | 3 (17.6%) |
| Both Inpatients and Outpatients | 6 (35.3%) |
| Community | 0 (0%) |
| Jail | 1 (5.8%) |
| **Pathogen** |  |
| MRSA or Other *Staphylococcus aureus* | 12 (70%) |
| *Enterococcus faecium* | 0 (0%) |
| *Escherichia coli* | 3 (17.6%) |
| *Pseudomonas aeruginosa* | 0 (0%) |
| *Klebsiella pneumoniae* | 0 (0%) |
| Enterobacterales | 1 (5.9%) |
| SSTI | 1 (5.9%) |
| UTI | 1 (5.9%) |
| **Funding type** |  |
| Academia only | 0 (0%) |
| Federal only | 3 (17.6%) |
| Industry only | 2 (11.7%) |
| Mixed | 3 (17.6%) |
| None | 4 (23.5%) |
| Not reported | 5 (29.4%) |
| **Reported directionality of effect measure, compared to White majority** |  |
| Higher risk | 4 (23.5%) |
| No significant difference | 5 (29.4%) |
| Lower risk | 3 (17.6%) |

Note: MRSA=Methicillin-resistant *S. aureus*. UTI=Urinary tract infection; SSTI=skin and soft tissue infection.

Compared to non-Hispanic White persons, two studies in the southern United States^49,62^ reported a higher risk of MRSA vs. MSSA infection among Hispanic patients and one reported a higher risk of MRSA colonization vs. no MRSA colonization among Hispanic adults.^59^ Meanwhile, one study reported a lower risk of MRSA vs. MSSA,^10^ four reported no difference between Hispanic and White persons for MRSA vs. MSSA,^21,26,32,33^ MRSA vs *S. aureus*, and MSSA vs no *S. aureus*, and four did not report an analysis comparing outcomes between Hispanic and White persons.^9,45,51,69^ Of the five studies that reported Enterobacterales infections in Hispanic patients,^25,46,60,70,71^ one study reported a higher risk of trimethoprim-sulfamethoxazole-resistant (TMP/SMX) versus TMP/SMX-susceptible *E. coli* infections,^25^ a San Francisco-based study reported no difference in ESBL-producing *E. coli* infections between races,^60^ and one did not report results for a comparison.^70^ Another study of 175 US hospitals compared CRE vs carbapenem-susceptible Enterobacterales infection rates between multiple races but did not report results for a comparison between Hispanic vs White persons.^71^ Pregnant women with and without antepartum pyelonephritis caused by *E. coli* or *Enterobacter* spp. were examined in one Texas study,^46^ but no difference between races was found.

One study reported on SSTI vs No SSTI. The authors found a lower risk of SSTI caused by S*. aureus*, *P. aeruginosa*, and *E. coli* in a mixed-aged population of Hispanic persons in metro Atlanta, compared to White persons.^62^

#### Asian vs. White persons

Asian persons were reported to either have a lower risk or no difference in risk for MRSA infection compared to other races in five studies, all of which were conducted in the USA.^21,33,45,56,62^ Two studies reported a lower risk of MRSA vs. MSSA infection or colonization in pediatric and mixed-age Asian populations as compared to other races,^45,62^ while three reported no significant difference between races for MRSA vs. MSSA infection or no *S. aureus* colonization.^21,33,56^

No difference was found between Asian and White persons in four studies that compared MSSA or *S. aureus* infection or colonization with no *S. aureus*.^33,47,56,62^ Two of these studies also reported data for SSTI vs no SSTI with one New Zealand reporting a higher risk in Asian patients compared to those of European descent,^47,62^ while a US study found a mixed age group of Asian individuals to be at a lower risk of SSTI than White individuals.^62^

Three studies reported data for Asian patients with *E. coli* infection, with varied results.^27,60,61^ Of these, one study of adult women in Philadelphia with fluoroquinolone-susceptible *E. coli* UTIs reported a higher risk of infections expressing higher-level resistance to levofloxacin (MIC >0.12) among Asian than White women,^61^ another study reported no difference between races, and a third (20) didn’t evaluate the comparison.^60^

#### Aboriginal, Māori, Pacific Islander vs. White persons

Six cohorts in seven publications reported results from Australia and New Zealand, where Aboriginals, Māori, and Pacific Islanders are minority populations.^18,24,40,47,66–68^ These minority groups were generally found to be at higher risk of MRSA and *S. aureus* colonization or infection, as well as SSTI.

Four of these studies described MRSA colonization and infection among these ethnic groups.^18,24,40,67^ Two studies from Australia that included inpatients with either SSTI or pediatric thoracic emphysema reported that Aboriginal and Torres Strait Islander children, but not Pacific Islander children, had as much as a four times higher risk of MRSA than MSSA infection relative to Caucasian or not Aboriginal and Torres Strait Islander children.^24,40^ From New Zealand, two studies reported that relative to persons identifying as Māori, Asian, or of European descent, Pacific Islander persons were at a higher risk of MRSA infection compared to either MSSA or no MRSA.^18,68^ One of these studies also reported that Pacific Islander children were almost three times more likely to have MRSA infections than European children.^68^

Pacific Islanders and Māori inpatient children in New Zealand were also reported to be at a higher risk of *S. aureus* vs no *S. aureus* infection relative to children of European descent in two studies.^66,68^ Hobbs 2018 did not find a difference in *S. aureus* vs. no *S. aureus* colonization between races but reported that Pacific Islander (RR: 1.44; 95% CI: 1.28, 1.62) and Māori persons (RR: 1.46; 95% CI: 1.30, 1.64) were at a higher risk of SSTI versus no SSTI.^47^

#### Other races

Conflicting results were reported in studies that aggregated non-White racial groups into a single category and which reported on other races. Two USA studies compared MRSA infection risks among White to all non-White patients, with one reporting higher risks for MRSA vs MSSA and MRSA vs no MSSA cases among non-White than White patients, while the other did not report a significant difference.^30,58^ Non-White vs White persons were also compared for differences in risk of colonization/infection with MSSA or *S. aureus* versus no *S. aureus,* with two studies reporting a higher risk of infection or colonization in non-White persons,^30,57^ while one did not find a significant difference.^58^ One study reported a higher risk of *S. aureus* colonization in non-Hispanic White participants than in other races.^50^

Results were mixed for three USA studies ^3,28,52^ that reported MRSA infections among Native Hawaiian, Pacific Islander, or Samoan persons and for four USA studies ^20,36,45,62^ that included a small percentage of American Indian, Alaskan Native, or Eskimo persons, or a mix of these minorities.

### Ethnicity

#### Hispanic vs non-Hispanic persons

Ten cohorts in twelve publications reported Hispanic as an ethnicity (**Table 4**).^8,17,20,22,27,29,30,35,42,50,63,65^ MRSA or *S. aureus* was reported in eight studies with conflicting results, *E. coli* was reported in three with two reporting higher risk in Hispanic persons, *K. pneumoniae*was reported in one, *P. aeruginosa* in one, SSTI in one, and UTI in one.

**Table 4:** Characteristics of studies that included Hispanic persons as an ethnicity.

|  | **Total (%)** |
| --- | --- |
| **N studies** | 10 in 12 publications |
| **Age group** |  |
| Pediatric only | 2 (20%) |
| Adults only | 4 (40%) |
| Mixed age groups | 4 (40%) |
| **N analyzed range** | 119- 1904807 |
| **Study duration** | 56 days 8 years |
| **% Male range (N=9)** | 12.5- 78% |
| Women only | 0 (0%) |
| **Study country** |  |
| Australia | 0 (0%) |
| Brazil | 0 (0%) |
| Israel | 0 (0%) |
| New Zealand | 0 (0%) |
| USA | 10 (100%) |
| **Outcome type** |  |
| Colonization | 1 (10%) |
| Infection | 9 (90%) |
| **Recruitment setting** |  |
| Inpatient | 3 (30%) |
| Outpatients | 4 (40%) |
| Both Inpatients and Outpatients | 2 (20%) |
| Community | 1 (10%) |
| Jail | 0 ( 0%) |
| **Pathogen** |  |
| MRSA or Other *Staphylococcus aureus* | 8 (80%) |
| *Enterococcus faecium* | 0 ( 0%) |
| *Escherichia coli* | 3 (30%) |
| *Pseudomonas aeruginosa* | 1 (10%) |
| *Klebsiella pneumoniae* | 1 (10%) |
| Enterobacterales | 0 ( 0%) |
| SSTI | 1 (10%) |
| UTI | 1 (10%) |
| **Funding type** |  |
| Academia only | 0 ( 0%) |
| Federal only | 4 (40%) |
| Industry only | 0 ( 0%) |
| Mixed | 5 (50%) |
| None | 0 ( 0%) |
| Not reported | 2 (20%) |
| **Reported directionality of effect measure, compared to White majority** | 5 (50%) |
| Higher risk | 5 (50%) |
| No significant difference | 2 (20%) |
| Lower risk |  |

Note: MRSA=Methicillin-resistant *S. aureus*. UTI=Urinary tract infection; SSTI=skin and soft tissue infection.

Compared to Non-Hispanic White persons, one metro Atlanta-based study ^20^ reported a higher risk of MRSA infection vs. controls (patients with an unintentional traumatic brain injury) among Hispanic children while another study ^30^ that compared Hispanic to non-Hispanic persons reported a higher risk of MSSA vs no MSSA infection in a mixed-aged population. Three studies ^17,30,50^ reported no significant difference between ethnicities with regards to risk of colonization or infection with MRSA and *S. aureus*, while two observational studies ^22,42^ reported a lower risk of MRSA vs. MSSA compared to non-Hispanics or non-Hispanic White persons. Three studies included Hispanic individuals but did not investigate this relationship.^29,35,63^

Three studies reported *E. coli* infections in Hispanic persons.^27,63,65^ One pediatric study in Tennessee reported that Hispanic children were at a higher risk than non-Hispanic children for *E. coli* UTIs that were non-susceptible to first-generation cephalosporin antibiotics.^65^ Another study ^27^ that reported on two adult cohorts recruited from an outpatient population in Northern California found that Hispanic adults compared to non-Hispanic adults were at a higher risk of MDR vs non-MDR *E. coli* infections, while a third study ^63^ did not report on the comparison for Hispanic vs non-Hispanic White persons for infections caused by *E. coli* versus other pathogens.

One Texas-based study that compared patients with and without SSTI reported a significantly lower risk of SSTI among non-English speaking Hispanic persons compared to non-Hispanic White persons.^8^ No significant difference was found between English-speaking Hispanic persons compared to non-Hispanic White persons.

A retrospective cohort study of 601 patients from San Antonio, Texas reported a higher number of CAP cases caused by *P. aeruginosa*vs other pathogens among non-Hispanic White (20%) than Hispanic adults (7%).^63^ This same study reported no difference among CAP caused by *K. pneumoniae* vs other pathogens between Hispanic (4%) and non-Hispanic White (5%) adults.

#### Bedouin vs. Jewish persons

Bedouin persons were generally reported to have either a higher or similar risk of infection or colonization with the pathogens of interest, relative to Jewish persons in Israeli studies.^19,34,39,41,44,54^ Two studies compared the ethnicity of children either colonized or infected with MRSA to those with either MSSA or no MRSA.^19,39^ Both studies found that Arab or Bedouin children, who are the ethnic minority in Israel, were at a significantly higher risk of MRSA compared to Jewish children.

Outpatient Bedouin children were reported to be at a significantly higher risk of quinolone-resistant vs susceptible UTI caused by *K. pneumoniae, P. aeruginosa,* or *E. coli* than Jewish children in one study.^41^ Another study examined antibiotic susceptibility patterns among various pathogens (*K. pneumoniae*, *Enterobacter* spp., and *E. coli*) causing inpatient UTIs among children and reported a higher risk of resistance to some antibiotics (e.g., ampicillin, TMP/SMX, cephalosporins) among Bedouin versus Jewish children, but no significant difference for other antibiotics (e.g., nitrofurantoin, meropenem, amikacin).^44^ Another study reported a higher risk of UTI caused by ESBL-producing vs non-ESBL-producing bacteria in Arab children than Jewish children.^54^ In a prospective cohort study of 223 adults with UTIs caused by *E. coli*, *P. aeruginosa*, or *Enterococcus faecium*, no difference in antibiotic susceptibility patterns was observed between Bedouin versus non-Bedouins adults.^34^

#### Middle Eastern vs. non-Middle Eastern Caucasian persons

One USA study of 214 patients reported a higher risk for ESBL-producing *E. coli* UTIs among people of Middle Eastern descent compared to Caucasian persons of non-Middle Eastern descent.^70^ One additional Australian study included a very small number of Middle Eastern patients but did not report results for a comparison.

## DISCUSSION

This scoping review found global evidence for racial and ethnic disparities in colonization/infection with priority bacterial pathogens that are increasingly AMR. While most studies we included were conducted in the USA, we also included studies from other countries, *i.e.*, Brazil, Israel, Australia, and New Zealand. In general, persons belonging to racial or ethnic minority groups within these countries – especially Black persons in the USA and Brazil, Aboriginal persons in Australia and New Zealand, and Arabs or Bedouins persons in Israel - were often at higher risk for colonization/infection with the pathogens of interest (especially MRSA) compared to majority groups, despite there being no biological basis for such differences. Our findings indicate that global efforts to equitably prevent, diagnose, and treat bacterial infections that are increasingly AMR will be challenging unless strategies that account for the SDOH that likely drive these disparities are considered.

It is widely acknowledged that racial and ethnic disparities in disease risks are likely underpinned by the SDOH. An individual’s position in the social hierarchy has historically been linked to mortality risks in most industrialized nations; those experiencing social exclusion tend to experience the worst health outcomes, but a social gradient has been documented for those with middle and even high incomes.^72^ In a scoping review that was conducted in parallel to this one, we identified several SES characteristics that were associated with higher risks for colonization/infection with the same organisms reported here, including low educational attainment, low income levels, lack of healthcare access, residential crowding, and high neighborhood-level deprivation.^15^ It is more biologically plausible that some or all of these characteristics drive differences in colonization/infection rates among minority groups. However, we note that at least historically, many studies did not examine race, ethnicity, and the SDOH in conjunction; of the 54 studies we included here, a substantial proportion only considered race or ethnicity as risk factors (60%) or considered them independently (*e.g.*, only in univariate models) from SES factors (33%). Therefore, it remains challenging to identify which SES factors may truly underlie the racial and ethnic differences we report here. Given that the influence of a particular SES factor on health depends on the society or social environment in which a person lives (*e.g.*, having a low income may be less harmful in a country with free access to healthcare, education, and other social services), we note that identifying meaningful SDOH is highly context-dependent.^72^ Our findings are not meant to guide clinical decision making, but may be helpful for identifying specific racial or ethnic groups that would especially benefit from targeted collection and investigation of the SDOH that are underlying differences in their health outcomes, with the ultimate goal of improving public health.

We identified several gaps that merit consideration in future studies. First, studies rarely included whether participants’ race and ethnicity were self-reported. Given well-known discrepancies between the race or ethnicity that an individual might identify with and what hospital staff note in the medical record,^73^ this information is critical for evaluating the validity of a study’s findings and poses a major limitation for interpreting the results we present here. We suggest that this information is required when studies seek to report racial or ethnic differences for any health outcome. We also noted that within the USA specifically, studies classified race and ethnicity using different categorizations, which made it challenging to compare findings. While these differences did not prevent us from extracting comparisons for the White majority versus minority groups, a more streamlined approach would be useful; updated federal recommendations have recently been published which should ameliorate this issue in the future.

Second, we noted very few studies included Indigenous populations, outside of studies from Australia and New Zealand. Indigenous populations are often poorly represented in research, which makes it challenging to design disease prevention strategies that are effective among these groups.^74^ During our study screening phase, we noted several studies that described colonization/infection with the pathogens of interest exclusively among Indigenous populations. Because these studies did not include a comparator group (e.g., a non-Indigenous population), they did not meet our inclusion criteria and were excluded. Consistent findings from Australia and New Zealand that Aboriginal populations were at higher risk of *S. aureus* and MRSA infections strongly support the need for further research among Indigenous and First Nations populations in the USA, Canada, and Central and South America, where there are sizeable populations.

Third, some studies excluded non-English speakers. Doing so, unfortunately, can lead to bias against a population that, because of language barriers, could harbor distinct risks of infection/colonization with pathogens that are commonly AMR.^27^ One study we included from Texas, USA explicitly compared non-English speakers to other races or ethnicities with regards to SSTI risk and reported a lower risk of SSTI in the non-English speaking population.^8^ This finding demonstrates that this demographic may have different risks and should be represented in studies when possible.

Finally, most studies we included focused on *S. aureus* and *E. coli*. Only one study each reported data on *K. pneumoniae*, *E. faecium* and Enterobacterales, and two studies included *P. aeruginosa*as a pathogen. Future studies should consider differences by race, ethnicity, and SES in colonization or community-acquired infection with these pathogens; systematic reviews and meta-analyses cannot be recommended presently due to insufficient data.

This scoping review had strengths. We systematically searched the literature in consultation with an information scientist (R.M.) and search terms were iteratively designed in consultation with experts in systematic reviews (N.N., E.A.), antimicrobial resistance and stewardship (M.L.N., S.D.), and clinical infectious diseases (S.D.). Screening and data extraction were conducted in duplicate to minimize subjectivity in data analysis. We included 54 studies comprising an analytical dataset of 3,377,156 persons. To the best of our knowledge, this is the first compilation of global evidence for racial and ethnic disparities in colonization and infection with priority bacterial pathogens that are increasingly AMR.

This study also had some limitations. First, we recognize that interpretations of race and ethnicity vary widely and thus umbrella terms like “Asian,” which were frequently employed by studies here, may change meaning from one region to another and intersect differently with the SDOH. We used terms employed by study authors and clarified study location when presenting results where possible, but acknowledge that these inherent differences may make comparisons between and even within countries more challenging. Second, the collection of race and ethnicity data is not routine and is even prohibited in some countries (e.g., France). This limited the representativeness of the studies we could include. Third, we excluded many studies because they did not report results stratified by race or ethnicity. We noted during our initial screening phase that most studies had enough information to report these data, but did not, likely because patients’ race and ethnicity was a covariate they controlled for but not an exposure of interest. In future publications, providing health outcomes stratified by race or ethnicity would be helpful.

## Conclusions

This scoping review, together with our concurrent review of the association between SES and colonization/infection with community-acquired pathogens, establishes the critical need to better delineate the underlying causes of disparities in colonization/infection with priority pathogens that are increasingly AMR. Several countries and regional institutions have already published recommendations to collect such data, including the United Kingdom and the WHO regional office for Europe,^75,76^ though implementation remains sporadic.^13^ We recommend that future studies focus on collecting self-reported race and ethnicity in conjunction with the SDOH, make a concerted effort to include non-English speakers and Indigenous populations in the Americas, and expand analyses beyond *S. aureus and E. coli* to other leading cause of AMR-associated deaths.^77^

## Supporting information

Suppplemental File

PRISMA Scoping Review Checklist

## ABBREVIATIONS

A: 

MR: Antimicrobial resistance

ATSI: Aboriginal and Torres Strait Islander

CA: Community-acquired

CA-AMR: Community-acquired antimicrobial resistance

CAP: Community-acquired pneumonia

CRE: Carbapenem-resistant Enterobacter ales

ESBL: Extended Spectrum Beta-Lactamase

ESKAPE: Enterococcus faecium, Staphylococcus aureus, Klebsiella pneumoniae, Acinetobacter baumannii, Pseudomonas aeruginosa, and Enterobacter spp.

MDR: Multi-drug resistant

MIC: Minimum inhibitory concentration

MRSA: Methicillin-resistant Staphylococcus aureus

MSSA: Methicillin-Sensitive Staphylococcus aureus

P. aeruginosa: Pseudomonas aeruginosa

S. aureus: Staphylococcus aureus

SES: Socioeconomic Status

SSTI: Skin and soft tissue infections

USA: United States of America

UTI: Urinary Tract Infection

## DECLARATIONS

### Authors’ contributions

MLN, CWC, and SD conceptualized the study. MLN, NN, and SD acquired the funding. RM, SD, NN, and CWC performed the literature search. SAB, EEA, NN, CWC, S Balaji, LM, SAA, SD, and MLN reviewed titles and abstracts, reviewed full texts, designed the data extraction template, and extracted data. EEA and SB prepared the tables and figures. EEA, RM, and MLN wrote the first draft of the manuscript. All authors reviewed and approved the manuscript.

### Availability of data and materials

Additional tables and figures are available in the supplementary material.

## Ethics approval and consent to participate

Not applicable.

## Consent for publication

Not applicable.

## Data Availability

All data produced in the present work are contained in the manuscript.

## Acknowledgements

Not applicable.

## REFERENCES

1 Einsiedel LJ, Fernandes LA, Woodman RJ. Racial disparities in infection-related mortality at Alice Springs Hospital, Central Australia, 2000--2005. Med J Aust 2008; 188: 568–71.

2 Centers for Disease Control and Prevention (CDC). Racial/ethnic disparities in diagnoses of HIV/AIDS--33 states, 2001-2004. MMWR Morb Mortal Wkly Rep 2006; 55: 121–5.

3 Centers for Disease Control and Prevention (CDC). Community-associated methicillin-resistant Staphylococcus aureus infections in Pacific Islanders--Hawaii, 2001-2003. MMWR Morb Mortal Wkly Rep 2004; 53: 767–70.

4 World Health Organization. Global Priority List of Antibiotic-Resistant Bacteria to Guide Research, Discovery, and Development of New Antibiotics. Geneva, Switzerland: WHO, 2017 https://www.who.int/news-room/fact-sheets/detail/antimicrobial-resistance.

5 Freedberg DE, Zhou MJ, Cohen ME, et al. Pathogen colonization of the gastrointestinal microbiome at intensive care unit admission and risk for subsequent death or infection. Intensive Care Med 2018; 44: 1203–11.

6 Huang SS, Septimus E, Kleinman K, et al. Chlorhexidine versus routine bathing to prevent multidrug-resistant organisms and all-cause bloodstream infections in general medical and surgical units (ABATE Infection trial): a cluster-randomised trial. Lancet 2019; 393: 1205–15.

7 Poudel AN, Zhu S, Cooper N, et al. The economic burden of antibiotic resistance: A systematic review and meta-analysis. PLoS One 2023; 18: e0285170.

8 Hemmige V, Arias CA, Pasalar S, Giordano TP. Skin and Soft Tissue Infection in People Living With Human Immunodeficiency Virus in a Large, Urban, Public Healthcare System in Houston, Texas, 2009-2014. Clin Infect Dis 2020; 70: 1985–92.

9 Bar-Meir M, Tan TQ. Staphylococcus aureus skin and soft tissue infections: can we anticipate the culture result? Clin Pediatr (Phila) 2010; 49: 432–8.

10 Hota B, Ellenbogen C, Hayden MK, Aroutcheva A, Rice TW, Weinstein RA. Community-associated methicillin-resistant Staphylococcus aureus skin and soft tissue infections at a public hospital: do public housing and incarceration amplify transmission? Arch Intern Med 2007; 167: 1026–33.

11 Williams DR, Priest N, Anderson NB. Understanding associations among race, socioeconomic status, and health: Patterns and prospects. Health Psychol 2016; 35: 407– 11.

12 Flanagin A, Frey T, Christiansen SL, AMA Manual of Style Committee. Updated Guidance on the Reporting of Race and Ethnicity in Medical and Science Journals. JAMA 2021; 326: 621–7.

13 He Z, Pfaff E, Guo SJ, et al. Enriching Real-world Data with Social Determinants of Health for Health Outcomes and Health Equity: Successes, Challenges, and Opportunities. Yearb Med Inform 2023; 32: 253–63.

14 Davis VH, Rodger L, Pinto AD. Collection and Use of Social Determinants of Health Data in Inpatient General Internal Medicine Wards: A Scoping Review. J Gen Intern Med 2023; 38: 480–9.

15 Blackmon SA, Avendano EE, Nirmala N, et al. Socioeconomic status and the risk for colonization or infection with priority bacterial pathogens: a global evidence map. In submission.

16 Covidence systematic review software. Veritas Health Innovation. Melbourne, Australia. Available at www.covidence.org.

17 Mainous AG, Hueston WJ, Everett CJ, Diaz VA. Nasal carriage of Staphylococcus aureus and methicillin-resistant S aureus in the United States, 2001-2002. Ann Fam Med 2006; 4: 132–7.

18 Abeysekera N, Wong S, Jackson B, Buchanan D, Heiss-Dunlop W, Mathy JA. Evolving Threat of Community Acquired Methicillin Resistant Staphylococcus aureus Upper Extremity Infections in the South Pacific: 2011-2015. J Hand Surg Asian Pac *Vol* 2019; 24: 129–37.

19 Adler A, Givon-Lavi N, Moses AE, Block C, Dagan R. Carriage of community-associated methicillin-resistant Staphylococcus aureus in a cohort of infants in southern Israel: risk factors and molecular features. J Clin Microbiol 2010; 48: 531–8.

20 Ali F, Immergluck LC, Leong T, et al. A Spatial Analysis of Health Disparities Associated with Antibiotic Resistant Infections in Children Living in Atlanta (2002-2010). EGEMS (Wash DC) 2019; 7: 50.

21 Berens P, Swaim L, Peterson B. Incidence of methicillin-resistant Staphylococcus aureus in postpartum breast abscesses. Breastfeed Med 2010; 5: 113–5.

22 Beresin GA, Wright JM, Rice GE, Jagai JS. Swine exposure and methicillin-resistant Staphylococcus aureus infection among hospitalized patients with skin and soft tissue infections in Illinois: A ZIP code-level analysis. Environ Res 2017; 159: 46–60.

23 Bes TM, Martins RR, Perdigão L, et al. Prevalence of methicillin-resistant Staphylococcus aureus colonization in individuals from the community in the city of Sao Paulo, Brazil. Rev Inst Med Trop Sao Paulo 2018; 60: e58.

24 Britton PN, Andresen DN. Paediatric community-associated Staphylococcus aureus: a retrospective cohort study. J Paediatr Child Health 2013; 49: 754–9.

25 Burman WJ, Breese PE, Murray BE, et al. Conventional and molecular epidemiology of trimethoprim-sulfamethoxazole resistance among urinary Escherichia coli isolates. Am J Med 2003; 115: 358–64.

26 Casey JA, Cosgrove SE, Stewart WF, Pollak J, Schwartz BS. A population-based study of the epidemiology and clinical features of methicillin-resistant Staphylococcus aureus infection in Pennsylvania, 2001-2010. Epidemiol Infect 2013; 141: 1166–79.

27 Casey JA, Rudolph KE, Robinson SC, et al. Sociodemographic Inequalities in Urinary Tract Infection in 2 Large California Health Systems. Open Forum Infect Dis 2021; 8: ofab276.

28 Castrodale LJ, Beller M, Gessner BD. Over-representation of Samoan/Pacific Islanders among patients with methicillin-resistant Staphylococcus aureus (MRSA) infections at a large family practice clinic in Anchorage, Alaska, 1996-2000. Alaska Med 2004; 46: 88–91.

29 Cohen AL, Shuler C, McAllister S, et al. Methamphetamine use and methicillin-resistant Staphylococcus aureus skin infections. Emerg Infect Dis 2007; 13: 1707–13.

30 Como-Sabetti KJ, Harriman KH, Fridkin SK, Jawahir SL, Lynfield R. Risk factors for community-associated Staphylococcus aureus infections: results from parallel studies including methicillin-resistant and methicillin-sensitive S. aureus compared to uninfected controls. Epidemiol Infect 2011; 139: 419–29.

31 Crum-Cianflone NF, Shadyab AH, Weintrob A, et al. Association of methicillin-resistant Staphylococcus aureus (MRSA) colonization with high-risk sexual behaviors in persons infected with human immunodeficiency virus (HIV). Medicine (Baltimore) 2011; 90: 379– 89.

32 David MZ, Mennella C, Mansour M, Boyle-Vavra S, Daum RS. Predominance of methicillin-resistant Staphylococcus aureus among pathogens causing skin and soft tissue infections in a large urban jail: risk factors and recurrence rates. J Clin Microbiol 2008; 46: 3222–7.

33 Duggal P, Naseri I, Sobol SE. The increased risk of community-acquired methicillin-resistant Staphylococcus aureus neck abscesses in young children. Laryngoscope 2011; 121: 51–5.

34 Elnasasra A, Alnsasra H, Smolyakov R, Riesenberg K, Nesher L. Ethnic Diversity and Increasing Resistance Patterns of Hospitalized Community-Acquired Urinary Tract Infections in Southern Israel: A Prospective Study. Isr Med Assoc J 2017; 19: 538–42.

35 Forcade NA, Parchman ML, Jorgensen JH, et al. Prevalence, severity, and treatment of community-acquired methicillin-resistant Staphylococcus aureus (CA-MRSA) skin and soft tissue infections in 10 medical clinics in Texas: a South Texas Ambulatory Research Network (STARNet) study. J Am Board Fam Med 2011; 24: 543–50.

36 Frei CR, Makos BR, Daniels KR, Oramasionwu CU. Emergence of community-acquired methicillin-resistant Staphylococcus aureus skin and soft tissue infections as a common cause of hospitalization in United States children. J Pediatr Surg 2010; 45: 1967–74.

37 Fridkin SK, Hageman JC, Morrison M, et al. Methicillin-resistant Staphylococcus aureus disease in three communities. N Engl J Med 2005; 352: 1436–44.

38 Fritz SA, Hogan PG, Hayek G, et al. Staphylococcus aureus colonization in children with community-associated Staphylococcus aureus skin infections and their household contacts. Arch Pediatr Adolesc Med 2012; 166: 551–7.

39 Galper E, Bdolah-Abram T, Megged O. Assessment of infections rate due to community-acquired Methicillin-resistant Staphylococcus aureus and evaluation of risk factors in the paediatric population. Acta Paediatr 2021; 110: 1579–84.

40 Gautam A, Wiseman GG, Goodman ML, et al. Paediatric thoracic empyema in the tropical North Queensland region of Australia: Epidemiological trends over a decade. J Paediatr Child Health 2018; 54: 735–40.

41 Gottesman B-S, Low M, Almog R, Chowers M. Quinolone Consumption by Mothers Increases Their Children’s Risk of Acquiring Quinolone-Resistant Bacteriuria. Clin Infect Dis 2020; 71: 532–8.

42 Graham PL, Lin SX, Larson EL. A U.S. population-based survey of Staphylococcus aureus colonization. Ann Intern Med 2006; 144: 318–25.

43 Gualandi N, Mu Y, Bamberg WM, et al. Racial Disparities in Invasive Methicillin-resistant Staphylococcus aureus Infections, 2005-2014. Clin Infect Dis 2018; 67: 1175–81.

44 Hain G, Goldbart A, Sagi O, Ben-Shimol S. High Rates of Antibiotic Nonsusceptibility in Gram-negative Urinary Tract Infection in Children With Risk Factors Occurring in the Preceding Month: Considerations for Choosing Empiric Treatment. Pediatr Infect Dis J 2021; 40: 639–44.

45 Hermos C, Shiau R, Hsiang M, Chambers H, Pan E. Epidemiology of community-associated methicillin-resistant Staphylococcus aureus in San Francisco children. J Pediatr Infect Dis 2015; 04: 247–59.

46 Hill JB, Sheffield JS, McIntire DD, Wendel GD. Acute pyelonephritis in pregnancy. Obstet Gynecol 2005; 105: 18–23.

47 Hobbs MR, Grant CC, Thomas MG, et al. Staphylococcus aureus colonisation and its relationship with skin and soft tissue infection in New Zealand children. Eur J Clin Microbiol Infect Dis 2018; 37: 2001–10.

48 Huppert JS, Bennett K, Kollar LM, Pattullo L, Mortensen JE. MRSA: rare in the vagina. J Pediatr Adolesc Gynecol 2011; 24: 315–6.

49 Immergluck LC, Leong T, Malhotra K, et al. Geographic surveillance of community associated MRSA infections in children using electronic health record data. BMC Infect Dis 2019; 19: 170.

50 Kuehnert MJ, Kruszon-Moran D, Hill HA, et al. Prevalence of Staphylococcus aureus nasal colonization in the United States, 2001-2002. J Infect Dis 2006; 193: 172–9.

51 Lee GC, Dallas SD, Wang Y, et al. Emerging multidrug resistance in community-associated Staphylococcus aureus involved in skin and soft tissue infections and nasal colonization. J Antimicrob Chemother 2017; 72: 2461–8.

52 Len KA, Bergert L, Patel S, Melish M, Kimata C, Erdem G. Community-acquired Staphylococcus aureus pneumonia among hospitalized children in Hawaii. Pediatr Pulmonol 2010; 45: 898–905.

53 Lipsky BA, Pecoraro RE, Chen MS, Koepsell TD. Factors affecting staphylococcal colonization among NIDDM outpatients. Diabetes Care 1987; 10: 483–6.

54 Megged O. Extended-spectrum β-lactamase-producing bacteria causing community-acquired urinary tract infections in children. Pediatr Nephrol 2014; 29: 1583–7.

55 Milstone AM, Carroll KC, Ross T, Shangraw KA, Perl TM. Community-associated methicillin-resistant Staphylococcus aureus strains in pediatric intensive care unit. Emerg Infect Dis 2010; 16: 647–55.

56 Morita JE, Fujioka RS, Tice AD, et al. Survey of methicillin-resistant Staphylococcus aureus (MRSA) carriage in healthy college students, Hawai’i. Hawaii Med J 2007; 66: 213–5.

57 Mork RL, Hogan PG, Muenks CE, et al. Longitudinal, strain-specific Staphylococcus aureus introduction and transmission events in households of children with community-associated meticillin-resistant S aureus skin and soft tissue infection: a prospective cohort study. Lancet Infect Dis 2020; 20: 188–98.

58 Nerby JM, Gorwitz R, Lesher L, et al. Risk factors for household transmission of community-associated methicillin-resistant Staphylococcus aureus. Pediatr Infect Dis J 2011; 30: 927–32.

59 Popovich KJ, Smith KY, Khawcharoenporn T, et al. Community-associated methicillin-resistant Staphylococcus aureus colonization in high-risk groups of HIV-infected patients. Clin Infect Dis 2012; 54: 1296–303.

60 Raphael E, Glymour MM, Chambers HF. Trends in prevalence of extended-spectrum beta-lactamase-producing Escherichia coli isolated from patients with community- and healthcare-associated bacteriuria: results from 2014 to 2020 in an urban safety-net healthcare system. Antimicrob Resist Infect Control 2021; 10: 118.

61 Rattanaumpawan P, Nachamkin I, Bilker WB, et al. Risk factors for ambulatory urinary tract infections caused by high-MIC fluoroquinolone-susceptible Escherichia coli in women: results from a large case-control study. J Antimicrob Chemother 2015; 70: 1547– 51.

62 Ray GT, Suaya JA, Baxter R. Incidence, microbiology, and patient characteristics of skin and soft-tissue infections in a U.S. population: a retrospective population-based study. BMC Infect Dis 2013; 13: 252.

63 Restrepo MI, Velez MI, Serna G, Anzueto A, Mortensen EM. Antimicrobial resistance in Hispanic patients hospitalized in San Antonio, TX with community-acquired pneumonia. Hosp Pract (1995) 2010; 38: 108–13.

64 See I, Wesson P, Gualandi N, et al. Socioeconomic Factors Explain Racial Disparities in Invasive Community-Associated Methicillin-Resistant Staphylococcus aureus Disease Rates. Clin Infect Dis 2017; 64: 597–604.

65 Stultz JS, Francis N, Ketron S, et al. Analysis of Community-Acquired Urinary Tract Infection Treatment in Pediatric Patients Requiring Hospitalization: Opportunity for Use of Narrower Spectrum Antibiotics. J Pharm Technol 2021; 37: 79–88.

66 Vogel AM, Borland A, van der Werf B, et al. Community-acquired invasive Staphylococcus aureus: Uncovering disparities and the burden of disease in Auckland children. J Paediatr Child Health 2020; 56: 244–51.

67 Williamson DA, Ritchie SR, Lennon D, et al. Increasing incidence and sociodemographic variation in community-onset Staphylococcus aureus skin and soft tissue infections in New Zealand children. Pediatr Infect Dis J 2013; 32: 923–5.

68 Williamson DA, Ritchie SR, Roberts SA, et al. Clinical and molecular epidemiology of community-onset invasive Staphylococcus aureus infection in New Zealand children. Epidemiol Infect 2014; 142: 1713–21.

69 Worley ML, Seif JM, Whigham AS, Mims JW, Shetty AK, Evans AK. Suppurative cervical lymphadenitis in infancy: microbiology and sociology. Clin Pediatr (Phila) 2015; 54: 629– 34.

70 Zhu FH, Rodado MP, Asmar BI, Salimnia H, Thomas R, Abdel-Haq N. Risk factors for community acquired urinary tract infections caused by extended spectrum β-lactamase (ESBL) producing Escherichia coli in children: a case control study. Infect Dis (Lond) 2019; 51: 802–9.

71 Zilberberg MD, Nathanson BH, Sulham K, Fan W, Shorr AF. Carbapenem resistance, inappropriate empiric treatment and outcomes among patients hospitalized with Enterobacteriaceae urinary tract infection, pneumonia and sepsis. BMC Infect Dis 2017; 17: 279.

72 Marmot MG. Understanding social inequalities in health. Perspect Biol Med 2003; 46: S9–23.

73 Magaña López M, Bevans M, Wehrlen L, Yang L, Wallen GR. Discrepancies in Race and Ethnicity Documentation: a Potential Barrier in Identifying Racial and Ethnic Disparities. J Racial Ethn Health Disparities 2016; 4: 812–8.

74 Vigil D, Sinaii N, Karp B. American Indian and Alaska Native Enrollment in Clinical Studies in the National Institutes of Health’s Intramural Research Program. Ethics Hum Res 2021; 43: 2–9.

75. Department of Health and Social Care, The Scottish Government, Welsh Government, Department for Environment, Food & Rural Affairs, Department of Health (Northern Ireland) and Department of Agriculture, Environment and Rural Affairs (Northern Ireland). UK 5-year action plan for antimicrobial resistance 2024 to 2029. United Kingdom, 2024 https://assets.publishing.service.gov.uk/media/664394d9993111924d9d3465/confronting-antimicrobial-resistance-2024-to-2029.pdf.

76 The socioeconomic drivers and impacts of Antimicrobial Resistance (AMR): Implications for policy and research. European Observatory on Health Systems and Policies, 2024 https://eurohealthobservatory.who.int/publications/i/the-socioeconomic-drivers-and-impacts-of-antimicrobial-resistance-(amr)-implications-for-policy-and-research.

77 GBD 2019 Antimicrobial Resistance Collaborators. Global mortality associated with 33 bacterial pathogens in 2019: a systematic analysis for the Global Burden of Disease Study 2019. Lancet 2022; 400: 2221–48.

