## Supplementary material for "Race, ethnicity, and risk for colonization and infection with key bacterial pathogens: a scoping review": Suppplemental File

**Supplementary Table S1.** Inclusion and exclusion criteria

| **Inclusion Criteria** | | **Exclusion Criteria** |
| --- | --- | --- |
| Study design | Observational studies | Case reports/study  Case series  Narrative Review |
| Study population | Any age group, gender, country, and health status | Studies in which data for race or ethnicity were not reported |
| Exposure | Any race or ethnic population of a country | Data stratified by country, region, or hospital rather than a racial/ethnic group |
| Comparator | Distinct racial/ethnic group from the same country | No distinct racial/ethnic group reported (e.g., all subjects are American Indian) |
| Outcomes | Community-acquired (e.g. outpatient, ERs, health clinics, ambulatory care, population-level surveillance) colonization or infection with:   - *Enterococcus faecium,* - *Staphylococcus aureus*, - *Klebsiella pneumoniae*, - *Acinetobacter baumannii*, - *Pseudomonas aeruginosa*, - *Enterobacter* species, - *Escherichia coli*, - Enterobacteriaceae, or Enterobacterales | - Hospital-acquired colonization or infection - Device-associated (e.g. catheter) infection - Non-bacterial infection or cause of infection unclear - Mixed pathogen infections if less than 50% of the pathogens were of interest - Community-acquisition or community-association based only on phenotype |

**Supplementary Table S2.** Ovid MEDLINE Search strategy

| 1 | Community-Acquired Infections/ |
| --- | --- |
| 2 | community acquired.mp. |
| 3 | community associated.mp. |
| 4 | community onset.mp. |
| 5 | Outpatients |
| 6 | outpatient$.mp. |
| 7 | Ambulatory Care/ |
| 8 | ambulatory.mp. |
| 9 | 1 or 2 or 3 or 4 or 5 or 6 or 7 or 8 |
| 10 | exp Socioeconomic Factors/ |
| 11 | socioeconomic$.mp. |
| 12 | socio-economic$.mp. |
| 13 | (standard$ adj2 living).mp. |
| 14 | social inequit$.mp. |
| 15 | social inequalit$.mp. |
| 16 | Health Status Disparities/ |
| 17 | Healthcare Disparities/ |
| 18 | (Health$ adj3 Disparit$).mp. |
| 19 | exp continental population groups/ or exp ethnic groups/ |
| 20 | (race$ or racial).mp. |
| 21 | ethnic$.mp. |
| 22 | 10 or 11 or 12 or 13 or 14 or 15 or 16 or 17 or 18 or 19 or 20 or 21 |
| 23 | Enterococcus faecium/ |
| 24 | exp Staphylococcus aureus/ |
| 25 | exp Staphylococcal Infections/ |
| 26 | Klebsiella pneumoniae/ |
| 27 | Acinetobacter baumannii/ |
| 28 | Pseudomonas aeruginosa/ |
| 29 | exp Enterobacter/ |
| 30 | exp Escherichia coli/ |
| 31 | exp Citrobacter/ |
| 32 | Enterobacteriaceae/ |
| 33 | Enterococcus faecium.mp. |
| 34 | Staphylococcus aureus.mp. |
| 35 | Klebsiella pneumoniae.mp. |
| 36 | Acinetobacter baumannii.mp. |
| 37 | Pseudomonas aeruginosa.mp. |
| 38 | Enterobacter.mp. |
| 39 | Escherichia coli.mp. |
| 40 | E coli.mp. |
| 41 | Citrobacter.mp. |
| 42 | gram-negative bacterial infections/ |
| 43 | exp enterobacteriaceae infections/ |
| 44 | exp moraxellaceae infections/ |
| 45 | Gram-Negative Bacteria/ |
| 46 | gram negative.mp. |
| 47 | Enterobacteriaceae.mp. |
| 48 | ESKAPE.mp. |
| 49 | 23 or 24 or 25 or 26 or 27 or 28 or 29 or 30 or 31 or 32 or 33 or 34 or 35 or 36 or 37 or 38 or 39 or 40 or 41 or 42 or 43 or 44 or 45 or 46 or 47 or 48 |
| 50 | 9 and 22 and 49 |

**Supplementary Table S3.** Individual characteristics of included studies

| **Author Year PMID** | **Country** | **Funding source** | **Study duration** | **Included Pathogens** | **Outcome** | **Recruitment method** | **Age group** | **N analyzed** | **% Male** | **Carriage or Infection Sites** | **Pathogen** |
| --- | --- | --- | --- | --- | --- | --- | --- | --- | --- | --- | --- |
| Abeysekera 2019 31035877 | New Zealand | NR | 5 years | *S. aureus*;  *P. aeruginosa*;  *E. coli* | Infection | Inpatient | Adult | 1,252 | 67.9 | SSTI | MRSA |
| Adler 2010 20007386 | Israel | NR | 10 months | *S. aureus* | Colonization | Community-based | Pediatric | 553 | NR | Colonization | MRSA |
| Ali 2019 31565665 | United States | Academia; Federal | 8 years | *S. aureus* | Infection | Inpatient; Outpatient | Pediatric | 39,371 | 60.1 | SSTI; uTBI | MRSA |
| Bar-Meir 2010 20118096 | United States | None | 18 months | *S. aureus* | Infection | Inpatient | Pediatric | 81 | 49 | SSTI | MRSA |
| Berens 2010 20113200 | United States | NR | 7 years | *S. aureus* | Infection | Inpatient | Adult | 33 | 0 | Breast abscess | MRSA |
| Beresin 2017 28772149 | United States | Federal; Non-profit | 3 years 7 months | *S. aureus* | Infection | Inpatient | Adult; Pediatric | 215,218 | 50.9 | SSTI | MRSA |
| Bes 2018 30365641 | Brazil | NR | 4 months | *S. aureus* | Colonization | Outpatient | Adult | 300 | 39 | Nasal colonization | MRSA |
| Britton 2013 23721234 | Australia | None | 1 year | *S. aureus* | Infection | Inpatient | Pediatric | 431 | 56.4 | SSTI | MRSA |
| Burman 2003 14553870 | United States | NR | 6 months | *E. coli* | Infection | Outpatient | Adult; Pediatric | 681 | 5.4 | UTI | *E. coli* |
| Casey 2013 22929058 | United States | Academia; Federal | 9 years total  (5 years with CA specified) | *S. aureus* | Infection | Inpatient; Outpatient | Adult; Pediatric | 8385 | 48.4 | SSTI | MRSA, SSTI |
| Casey 2021 34189179 | United States | Federal | 3 years | *E. coli* | Infection | Outpatient | Adult | 1,904,807 | 12.5 | UTI | *E. coli*, UTI |
| Castrodale 2004 15999910 | United States | NR | 3 years 4 months | *S. aureus* | Infection | Outpatient | Adult; Pediatric | 63 | NR | SSTI | MRSA, *S. aureus* |
| CentersforDisease 2004 15329653 | United States | NR | 2 years | *S. aureus* | Infection | Inpatient; Outpatient | Adult; Pediatric | 346 | 61 | SSTI | MRSA |
| Cohen 2007 18217555 | United States | NR | 56 days | *S. aureus* | Infection | Outpatient | Adult; Pediatric | 403 | 42.18 | SSTI | MRSA, MSSA |
| Como-Sabetti 2011 20513251 | United States | Federal | 3 years | *S. aureus* | Infection | Inpatient; Outpatient | Adult; Pediatric | 588 | 41 | SSTI; Non-SSTI/non-invasive (ear, eye, urine), and invasive (blood, joint, bone) | MRSA, MSSA |
| Crum-Cianflone 2011 22033452 | United States | Federal | 3 years | *S. aureus* | Colonization | Outpatient | Adult | 550 | 93 | SSTI | MRSA |
| David 2008 18685002 | United States | Industry; Federal | 18 months | *S. aureus* | Infection | Jail | Adult | 283 | 80 | SSTI | MRSA |
| Duggal 2011 21120830 | United States | None | 5 years | *S. aureus* | Infection | Inpatient | Pediatric | 136 | 43.22 | SSTI | MRSA, MSSA |
| Elnasasra 2017 28971635 | Israel | NR | 4 months | *Enterococcus faecium;*  *K. pneumoniae;*  *P. aeruginosa;*  *E. coli* | Infection | Inpatient | Adult | 223 | 29.1 | UTI | *E. coli*, *P. aeruginosa* |
| Forcade 2011 21900437 | United States | Federal | 1 year | *S. aureus* | Infection | Outpatient | Adult | 119 | 51 | SSTI | MRSA |
| Frei 2010 20920714 | United States | Academia | 11 years | *S. aureus* | Infection | Inpatient | Pediatric | 616,375 | 55 | SSTI | MRSA, *S. aureus* |
| Fridkin 2005 15814879 | United States | Federal | 12, 18, 24 months | *S. aureus* | Infection | Inpatient | Adult; Pediatric | 1,647 | NR | SSTI; bacteremia, septic arthritis, osteomyelitis, in wounds | MRSA |
| Fritz 2008 18519477 | United States | Federal | 8 months | *S. aureus* | Colonization | Outpatient | Pediatric | 1,300 | 51.6 | SSTI | MRSA, MSSA |
| Galper 2021 33249634 | Israel | NR | 12 years | *S. aureus* | Infection | Inpatient; Outpatient | Pediatric | 620 | 57.6 | SSTI | MRSA |
| Gautam 2018 29442395 | Australia | NR | 10 years | *S. aureus* | Infection | Inpatient | Pediatric | 123 | 58.5 | SSTI; Paediatric thoracic empyema | MRSA, MSSA |
| Gottesman 2020 31504346 | Israel | Academia | 7.5 years | *K. pneumoniae;*  *P. aeruginosa;*  *E. coli* | Infection | Outpatient | Pediatric | 40,204 | 6.2 | UTI | mixed |
| Graham 2006 16520472 | United States | Federal | 1 year | *S. aureus* | Colonization | Community-based | Adult; Pediatric | 9,622 | 48.7 | Nasal colonization | MRSA, *S. aureus* |
| Gualandi 2018 29659728 | United States | Federal | 10 years | *S. aureus* | Infection | Inpatient; Outpatient | Adult; Pediatric | 43,202 | NR | Invasive MRSA infection | MRSA |
| Hain 2021 33872277 | Israel | None | 1 year | *K. pneumoniae;*  *Enterobacter* species*;*  *E. coli* | Infection | Inpatient | Pediatric | 744 | 30% | UTI | Mixed |
| Hemmige 2020 31209457 | United States | Academia ; Federal | 6 years | *S. aureus* | Infection | Outpatient | Adult | 8,597 | 68.2 | SSTI | SSTI |
| Hermos 2009 None | United States | NR | 6 months | *S. aureus* | Infection | Inpatient; Outpatient | Pediatric | 170 | 53.5 | SSTI | MRSA |
| Hill 2005 15625136 | United States | NR | 2 years | *Enterobacter* species*;*  *E. coli* | Infection | Inpatient | Adult | 32,282 | 0 | Antepartum pyelonephritis | UTI |
| Hobbs 2018 30066280 | New Zealand | Academia; Industry; Federal; Non-profit | 4.5 years | *S. aureus* | Colonization | Community-based | Pediatric | 5,126 | 51.3 | SSTI; S. aureus colonization | *S. aureus*, SSTI |
| Hota 2007 17533205 | United States | None | 5 years and 8 months | *S. aureus* | Infection | Inpatient; Outpatient | Adult | 1,222 | NR | SSTI | MRSA |
| Huppert 2011 21872776 | United States | NR | 5 months | *S. aureus* | Colonization | Outpatient | Adult; Pediatric | 315 | 0 | Vaginal S. aureus colonization | *S. aureus* |
| Immergluck 2019 30777016 | United States | Academia; Federal; Non-profit | 9 years | *S. aureus* | Infection | Inpatient; Outpatient | Pediatric | 10,642 | 52.1 | SSTI | MRSA |
| Kuehnert 2006 16362880 | United States | Federal | 2 years | *S. aureus* | Colonization | Community-based | Adult; Pediatric | 9,622 | 48.7 | Nasal Colonization | MRSA, *S. aureus* |
| Lee 2017 28859442 | United States | Federal | 3 months | *S. aureus* | Infection | Outpatient | Adult | 111 | 50.45 | SSTI | MDR *S. aureus* |
| Len 2010 20632405 | United States | None | 12 years | *S. aureus* | Infection | Inpatient | Pediatric | 38 | 47 | SSTI | MRSA |
| Lipsky 1987 3622205 | United States | Federal | 6 months | *S. aureus* | Colonization | Outpatient | Adult | 103 | 98.1 | Carriage from nose and skin | *S. aureus* |
| Mainous III 2006 16569716 | United States | Federal; Non-profit | 2 years | *S. aureus* | Colonization | Community-based | Adult; Pediatric | 9,622 | NR | Nasal carriage | *S. aureus* |
| Megged 2014 24705795 | Israel | NR | 10 years | *K. pneumoniae;*  *Enterobacter species;*  *E. coli;* Enterobacteriaceae/Enterobacterales | Infection | Inpatient | Pediatric | 160 | 23.1 | UTI | UTI |
| Milstone 2010 20350379 | United States | Academia; Federal; Non-profit | 15 months | *S. aureus* | Colonization | Inpatient | Pediatric | 1,210 | 55 | Colonization | MRSA |
| Morita 2007 17941374 | United States | NR | "During the spring semester" | *S. aureus* | Colonization | Community-based | Adult | 100 | 29 | Colonization | MRSA, MSSA |
| Mork 2020 31784369 | United States | Federal | 3 years | *S. aureus* | Colonization | Community-based | Adult; Pediatric | 692 | 47.1 | Colonization | *S. aureus* |
| Nerby 2011 21617572 | United States | Federal | 1 year and 9 months | *S. aureus* | Infection | Inpatient; Outpatient | Pediatric | 232 | NR | NR | MRSA |
| Popovich 2012 22354926 | United States | Federal | 6 months | *S. aureus* | Colonization | Outpatient | Adult | 601 | 42 | Colonization | MRSA |
| Raphael 2021 34380549 | United States | Federal | 6 years 3 months | *E. coli* | Infection | Inpatient; Outpatient | Adult; Pediatric | 5,576 | 15 | Bacteriuria | *E. coli* |
| Rattanaumpawan 2015 25630645 | United States | Federal; Other: Pennsylvania Department of Health | 3 years | *E. coli* | Infection | Outpatient | Adult | 2,001 | 0 | UTI | *E. coli* |
| Ray 2013 23721377 | United States | Industry | 3 years | *S. aureus* | Infection | Inpatient; Outpatient | Adult; Pediatric | 376,262 | 47 | SSTI | MRSA, *S. aureus*, SSTI |
| Restrepo 2010 21068534 | United States | Academia; Federal; Non-profit | 4 years | *S. aureus;*  *K. pneumoniae;*  *P. aeruginosa;*  *E. coli* | Infection | Inpatient | Adult | 601 | 78 | CAP | MRSA, MSSA, *E. coli, K. pneumoniae, P. aeruginosa* |
| See 2017 28362911 | United States | Federal | 3 years | *S. aureus* | Infection | Inpatient; Outpatient | Adult | 2,521 | 63.5 | NR | MRSA |
| Stultz 2021 34752558 | United States | Other: None | 3 years | *E. coli* | Infection | Inpatient | Pediatric | 320 | 30 | UTI | *E. coli* |
| Vogel 2020 31355978 | New Zealand | Academia | 5 years | *S. aureus* | Infection | Inpatient | Pediatric | 295 | 68.47 | Invasive S. aureus infection | *S. aureus* |
| Williamson 2013 23518822 | New Zealand | Internal funding | 4 years | *S. aureus* | Infection | Inpatient | Pediatric | 1,860 | 53 | SSTI | MRSA |
| Williamson 2014 24534254 | New Zealand | Academia; Federal | 4 years | *S. aureus* | Infection | Inpatient | Pediatric | 163 | 56.4 | SSTI; Musculoskeletal, respiratory, endovascular, central nervous system, bacteremia | *S. aureus* |
| Worley 2015 25972051 | United States | None | 10 years | *S. aureus* | Infection | Inpatient | Pediatric | 76 | 43 | SSTI | MRSA |
| Zhu 2019 31429616 | United States | NR | 5 years | *E.coli* | Infection | Inpatient | Pediatric | 214 | 14.5 | UTI | *E. coli* |
| Zilberberg 2017 28415969 | United States | Industry | 5 years | *K. pneumoniae;*  *E. coli;* Enterobacteriaceae/Enterobacterales | Infection | Inpatient | Adult | 40,137 | 42.1 | CAP; UTI; Sepsis, Respiratory failure | CRE |

Note: CAP=Community-acquired pneumonia; CRE=Carbapenem-resistant Enterobacterales; MRSA=Methicillin-susceptible S. aureus; MRSA=Methicillin-resistant S. aureus; NR= Not reported; SSTI=Skin and soft tissue infection, UTI=Urinary tract infection.

Figures

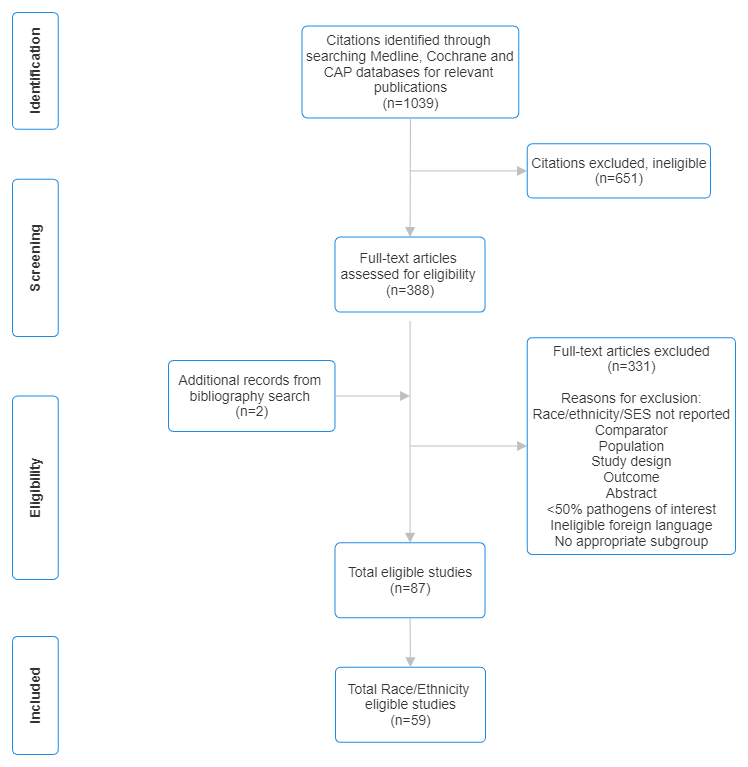

**Figure S1.** Study inclusion schema.
